# A survey to explore new markers of achievement to assess and monitor gender equity in an NIHR Biomedical Research Centre: A two-factor model

**DOI:** 10.1101/2020.02.04.20020347

**Authors:** Lorna R Henderson, Syed Ghulam Sarwar Shah, Pavel V Ovseiko, Rinita Dam, Alastair M Buchan, Helen McShane, Vasiliki Kiparoglou

## Abstract

**Background:** The underrepresentation of women in academic medicine at senior level and in leadership positions is well documented. In the United Kingdom, the National Institute for Health Research (NIHR) announced that eligibility for funding for Biomedical Research Centres (BRCs) required at least Silver award status of the Athena SWAN Charter. However, the evidence base for monitoring gender equity (GE) in BRCs is underdeveloped.

**Methods:** An exploratory online survey distributed to an entire population of NIHR Oxford BRC affiliates (N=683) who ranked the importance of 13 markers of GE on a five point Likert scale. Data were summarised using frequencies and descriptive statistics. Interrelationships between the markers and underlying latent dimensions (factors) were determined by exploratory and confirmatory factor analyses. Thematic analysis was used to analyse open-ended comments.

**Results:** The response rate was 36% (243 respondents). Respondents were more frequently female (55%, n=133), aged 41-50 years (33%, n=81), investigators (33%, n=81) and had been affiliated with the BRC for 2-7 years (39.5%, n=96). Participants ranked BRC senior leadership roles and organisational policies on gender equity, as very important, 58% (n=141) and 57% (n=139) respectively. The top two markers ranked as very important by female participants were organisational policies (64.7%, n=86/133) and recruitment and retention (60.9%, n=81/133), whereas male participants ranked leadership development (52.1%, n=50/96) and BRC senior leadership roles (50%, n=48/96) as being very important. The factor analyses showed two distinct latent dimensions: organisational markers and individual markers of GE in BRCs. Open ended comments suggested three key areas of actions: monitoring and benchmarking, organisational support for those with childcare responsibilities, and leadership and Institutional support for GE.

**Conclusions:** The findings suggest a two-factor model of markers of achievement for GE with organisational and individual dimensions. Implementation and sustainability of gender equity requires commitment at senior leadership and organisational policy level.

## Introduction

Underutilisation of female talent and potential in academic medicine, particularly at senior levels and leadership roles, as well as the health workforce more broadly is well documented [1–4]. This has been referred to as a “leaky talent pipeline” [5]. In 2011 the challenge to address gender equity (GE) in medical schools was linked directly to funding eligibility for Biomedical Research Centres (BRCs) in England. The UK Department of Health’s Chief Medical Officer announced that the National Institute for Health Research (NIHR) would not expect to shortlist any National Health Service (NHS) /University partnership for NIHR BRC designation and funding: “where the academic partner (generally the Medical School/Faculty of Medicine) has not achieved at least a Silver Award of the Athena SWAN Charter for Women in Science” [6].

### Athena SWAN Charter

The Athena SWAN charter advances women’s careers in higher education and research in terms of representation, progression of students into academia, journey through career milestones and working environment [7]. The higher education institutions (HEIs) can be awarded Bronze, Silver or Gold Athena SWAN award, based on their action plans, achievements and impact in advancing gender equity [7]. Athena SWAN awards are useful markers of GE achievement in higher education but they were not specifically designed for translational research organisations (TROs) such as NIHR BRCs, which are partnerships between UK’s leading NHS organisations and universities [8]. Furthermore the majority of GE research has focussed on perceptions of the Athena SWAN Charter in higher education settings [9–13]. A recent major evaluation of the Athena SWAN Charter did not examine Athena SWAN in the context of NIHR BRCs [9]. New measures to accelerate women’s advancement and leadership specific to NIHR BRCs have also been recommended [14]. This study aims to identify new markers of achievement for assessing and monitoring GE in NIHR BRCs.

## Methods

In this study, we adopt the UNESCO definition of gender equity, which refers to: “fairness of treatment for women and men, according to their respective needs. This may include equal treatment or treatment that is different but which is considered equivalent in terms of rights, benefits, obligations and opportunities” [15].

### Study design, participants and setting

The study design utilised a cross sectional study using an online questionnaire via SurveyMonkey® [16]. The participants included all researchers and staff affiliated with the NIHR Oxford BRC. The total sample (N=683) comprised all NHS consultants and university clinical academics (i.e., NIHR investigators and NIHR senior investigators); administrative and support staff (i.e., NIHR associates); academic and clinical trainees (i.e., NIHR academy members); patient and public involvement representatives, industry managers and leaders (including the most senior executive and non-executive committees in the NIHR Oxford BRC) funded/supported by the NIHR Oxford BRC (herewith referred to as BRC affiliates). These affiliates were selected because of their involvement in translational research, administration, support and leadership of the BRC. Names and contact details of all affiliates were extracted from the BRC’s internal databases. To ensure accuracy, all BRC theme managers were also contacted and asked to provide up to date email addresses of affiliates within their respective themes.

### Development of the questionnaire

We developed a survey questionnaire that comprised two parts. The first part of the survey comprised quantitative questions about thirteen markers of achievement of GE in BRCs and additional open ended questions asking respondents to add any other comments on measuring GE in BRCs. We identified markers from the literature and via qualitative analysis of face to face interviews (n=16) with women affiliated to the BRC as part of the larger project (Henderson et al, unpublished manuscript). In the first part of the survey, participants were asked to rank the importance of 13 markers of GE on a five point Likert scale: Very important” (score 5), “Important” (score 4), “Neutral” (score 3), “Not important” (score 2) and “Not at all important” (score 1).

The second part of the questionnaire asked for participants’ demographic characteristics i.e., age, gender; current role in the NIHR Oxford BRC and how long they had been affiliated to the BRC. Taking into account the diverse identities of women and men and based on the University of Oxford staff survey categories we did not use a binary sex indicator for gender but expanded it based on guidance from the University of Oxford’s equality and diversity team to offer the option to self-describe or not report.

### Piloting of the questionnaire

The questionnaire was piloted in two phases. In the first phase, it was piloted in face-to-face interviews with potential participants (n=10) to ensure it was easily understood and met the purpose of what it was intended to measure. In the second phase, it was tested via email to a small sample (n=16) from the population of interest to assess readability and clarity of the items in terms of consistency and appropriateness of interpretations from participants. Following the piloting, a few minor changes were made in wording and formatting prior to the main survey study.

### Administration of survey

The survey was conducted from May to July 2019. The NIHR Oxford BRC’s Chief Operating Officer sent an email via SurveyMonkey® with a web link to the anonymous online survey to BRC affiliates (N=683) informing them about the survey. The clinical research manager of the NIHR Oxford BRC also sent an email to all BRC theme liaisons (theme managers) to inform their theme members i.e., theme leaders, researchers and supporting staff about the survey. Up to 3 automated email reminders over the 6 weeks were sent via SurveyMonkey® to the participants who had not yet completed or had partially completed the survey.

### Data analysis

Online data from SurveyMonkey® was downloaded in SPSS and Microsoft Excel spreadsheet formats. We analysed data using frequencies of participants’ demographic characteristics and the descriptive statistics of scores of the importance of 13 markers of GE in BRCs. We used the Mann-Whitney U test to determine statistical differences in ranking the importance of GE markers by participants’ gender (male and female) and the Kruskall-Wallis H test to evaluate differences in ranking of the markers by the participants’ age, role and duration of affiliation to the BRC. Thereafter, data on participants’ scores of the importance of 13 markers of GE in BRCs were analysed using the exploratory and confirmatory factor analyses as described below.

#### Exploratory factor analysis

We determined the interrelationships between the markers and underlying latent dimensions (factors) by exploratory factor analysis (EFA) [17]. The EFA was run to extracting the latent factors (dimensions) covered in the measured 13 markers of GE. For the EFA, we used Principal Component Analysis (PCA) as a factor-extraction method, the Varimax with Kaiser Normalization as a rotation method and the Kaiser’s Eigen values > 1 (EVG1) criterion and breaks in the scree plot for determining the number of latent factors [18]. We applied minimum communalities ≥ 0.50, with no cross loadings ≥0.45 on more than one latent factor [18] and the minimum acceptable factor loading as 0.50 on only one factor [17]. Our sample size was 243 and the participant-to-variable ratio was 18:1, which was higher than the minimum acceptable participant-to-variable ratio of 10:1 [17].

#### Confirmatory factor analysis

Subsequent to the EFA, we ran the confirmatory factor analysis (CFA) [19]. The internal consistency of latent dimensions identified in the EFA was checked by running scale reliabilities using the Cronbach’s alpha coefficient [20]. The measurement model identified in CFA was checked for convergent and discriminant validity by calculating the Average Variance Extracted (AVE) as suggested [17,21].

Participants’ ratings of GE markers were positively skewed, which was reduced by log transformation prior to running EFA and CFA [19]. All statistical analyses were undertaken using IBM SPSS Statistics for Windows, version 25.0 (IBM Corp INC: Armonk, NY) except the CFA for which we used the IBM SPSS AMOS for windows, version 26.0 (IBM Corp INC: Armonk, NY).

#### Qualitative analysis

The open ended comments were analysed by LH, RD and SGSS. The comments were initially coded in Microsoft® Office; and NVIVO software [22] was used to organise data further. Thematic analysis was used to interpret the data and develop a coding tree [23]. The researchers discussed areas of agreement concerning coding of themes and addressed areas of disagreement. Through an iterative process the number of topics was consolidated.

### Ethics

The study was reviewed by the Oxford University Medical Sciences Inter-divisional Research Ethics Committee and the University of Oxford Clinical Trials and Research Governance office who determined that the study was exempt from full ethical review. The information sheet provided on the first page of the online survey informed participants that their participation in the survey was voluntary and they could withdraw at any time. They were also informed that their data and responses provided in the survey would be held securely, confidential, processed and reported anonymously and in aggregated format. Participants were informed that ‘if you do not wish to complete the survey, please click on ‘No, I do not consent’ and then the survey will be aborted’. Consequently, only those participants who gave their online informed consent by clicking the option ‘Yes, I consent’ were able to complete the survey via SurveyMonkey®.

## Results

### Response rate

The survey was completed by 277 out of 683 participants invited; however, 34 responses were ineligible for inclusion as they provided partial or missing data; hence, they were removed from the sample and data analysis. One participant did not consent and opted out of the survey. Therefore, our final sample comprised 243 respondents and the effective response rate was 36%.

### Demographic characteristics of participants

Table 1 shows that the majority of respondents were female (55%, n=133), aged 41-50 years (33.3%, n=81), investigators e.g. principal, co and chief investigators (33.3%, n=81) and those who had been affiliated with the BRC for 2-7 years (39.5%, n=96).

**Table 1.** Socio-demographic characteristics of survey respondents.

| **Socio-demographic characteristics** | **Frequency** | **Percentage** |
| --- | --- | --- |
| <u>Gender</u> |  |  |
| Female | 133 | 54.7 |
| Male | 96 | 39.5 |
| Prefer to self describe | 3 | 1.2 |
| Prefer not to say | 10 | 4.1 |
| Missing data | 1 | 0.4 |
| <u>Age (years)</u> |  |  |
| 18-30 | 21 | 8.6 |
| 31-40 | 60 | 27.4 |
| 41-50 | 81 | 33.3 |
| 51-60 | 52 | 21.4 |
| 61+ | 17 | 7.0 |
| Prefer not to say | 11 | 4.5 |
| Missing data | 1 | 0.4 |
| <u>Staff category / Role in the BRC</u> |  |  |
| Investigators (e.g. PI/co-PI/CI) | 81 | 33.3 |
| Research Associates (e.g. Researchers and research fellows) | 67 | 27.6 |
| Admin/technical/Professional/Support Associates | 59 | 24.3 |
| Trainees/PhD students | 3 | 1.2 |
| Prefer not to say | 9 | 3.7 |
| Other | 21 | 8.6 |
| Missing data | 3 | 1.2 |
| <u>Duration of affiliation with the BRC</u> |  |  |
| Up to 2 years | 86 | 35.4 |
| 3-7years | 96 | 39.5 |
| More than 7 years | 52 | 21.4 |
| Prefer not to say | 6 | 2.5 |
| Missing data | 3 | 1.2 |

### Ranking of markers of achievement for gender equity

Table 2 presents participants’ rankings of the importance of 13 markers of GE. The top two markers with the highest overall mean rankings were BRC senior leadership roles (mean = 4.43, standard deviation (SD) = 0.80) and organisational policies on gender equity (mean = 4.40, SD = 0.85).

**Table 2.** Participants ranking* of markers of achievement of gender equity in biomedical research centres (N=243).

| Markers of achievement of gender equity | Descriptive statistics |  |  |  |  | Differences in rating by gender<sup>a</sup> |  |  |  |  |
| --- | --- | --- | --- | --- | --- | --- | --- | --- | --- | --- |
|  | Mean | Standard Deviation | Median | Mode | Percentiles |  |  | Mann-Whitney U | Z score | Asymp. Sig. (2-tailed) |
|  |  |  |  |  | 25 | 50 | 75 |  |  |  |
| 1. BRC senior leadership roles: e.g. Director, Steering Committee Member, Theme leader & Co-lead. | 4.43 | 0.80 | 5 | 5 | 4 | 5 | 5 | 5492.00 | -2.055 | 0.040 |
| 2. Leadership development: e.g. Gender-sensitive leadership programmes, succession plans. | 4.34 | 0.91 | 5 | 5 | 4 | 5 | 5 | 6128.00 | -0.582 | 0.561 |
| 3. BRC staff category: e.g. Principal Investigator, Researchers, Trainees and Admin & Support staff. | 4.33 | 0.87 | 5 | 5 | 4 | 5 | 5 | 5471.00 | -2.053 | 0.040 |
| 4. Recruitment & retention: e.g. Number of Staff recruited and promoted. | 4.34 | 0.86 | 5 | 5 | 4 | 5 | 5 | 5211.00 | -2.651 | 0.008 |
| 5. BRC funding: e.g. Distribution by Theme, Gender and Role. | 4.13 | 0.96 | 4 | 5 | 4 | 4 | 5 | 5335.00 | -2.278 | 0.023 |
| 6. External grant funding: e.g. Total amount, Role on the grant, Number of grants and Success rate. | 4.09 | 0.94 | 4 | 5 | 4 | 4 | 5 | 5444.00 | -2.032 | 0.042 |
| 7. Esteem indicators: e.g. NIHR Senior Investigators, Funding panel membership, Invited plenary speakers, Fellowships of learned societies, Honours and Awards. | 4.25 | 0.91 | 4 | 5 | 4 | 4 | 5 | 5522.00 | -1.912 | 0.056 |
| 8. Publications: e.g. Authorship (First / Corresponding / Senior author) and Type of Publication (Journal articles and Conference papers). | 4.09 | 0.97 | 4 | 5 | 4 | 4 | 5 | 5483.00 | -1.955 | 0.051 |
| 9. Intellectual property: e.g. Number of Patents, Licenses and Spinouts. | 3.90 | 1.06 | 4 | 5 | 3 | 4 | 5 | 5505.50 | -1.867 | 0.062 |
| 10. Collaboration with industry: e.g. Board membership, Joint grants and Advisory roles (non-executive directorships). | 3.99 | 1.00 | 4 | 4 | 3 | 4 | 5 | 5434.00 | -2.045 | 0.041 |
| 11. Patient and public involvement: e.g. Representative Number of Men and Women Speakers and Participants. | 4.19 | 0.90 | 4 | 5 | 4 | 4 | 5 | 5536.00 | -1.856 | 0.063 |
| 12. Organisational policies on gender equity: e.g. Personal Development Training, Mentoring, Sponsorship and Career Development. | 4.40 | 0.85 | 5 | 5 | 4 | 5 | 5 | 5462.00 | -2.115 | 0.034 |
| 13. Organisational Targets: e.g. Creating BRC targets for Gender Equity. | 4.21 | 0.98 | 4 | 5 | 4 | 4 | 5 | 5375.00 | -2.230 | 0.026 |

\*Scores: 1 = Not important at all, 2 = Not important, 3 = Neutral, 4 = Important, 5 = Very Important. a. Grouping Variable: Gender

When scores for all participants were combined, then the majority (58%, n=141) of participants scored BRC senior leadership roles as a very important marker of GE followed by organisational policies on GE ranked as the second highest very important marker by 57.2% (n=139) of participants (Fig 1). Whereas the collaboration with industry and Intellectual property emerged as the last and second last very important markers of GE in BRCs reported by 35.4% (n=86) and 35.8% (n=87) of participants respectively (Fig 1).

**Fig 1.**
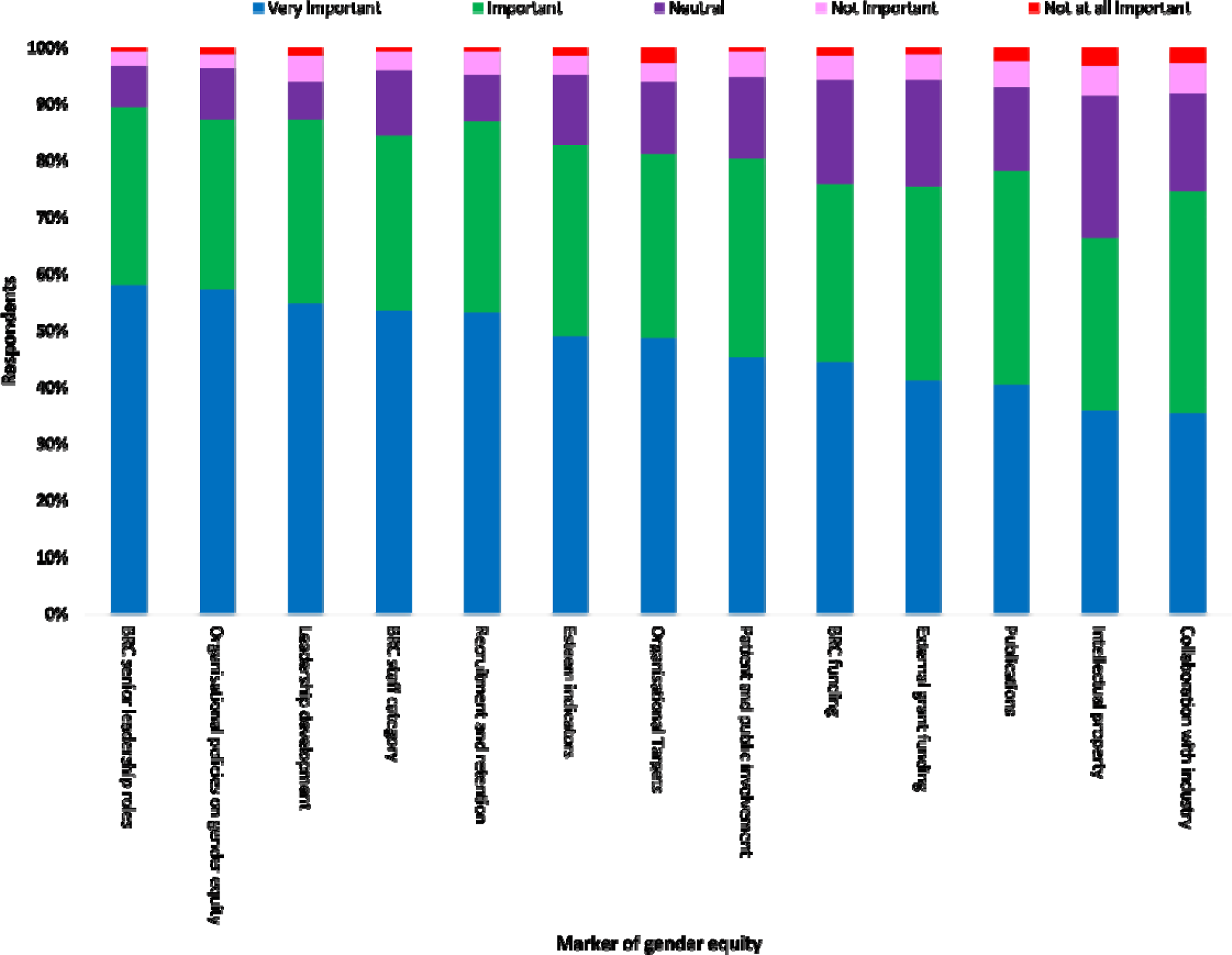
Importance of markers of gender equity in BRCs by all respondents.

There were statistically significant differences in the mean rankings by male and female participants for eight markers. These markers were BRC senior leadership roles (U=5492 p=0.040), BRC staff category (U=5471, p=0.040), recruitment and retention, (U=5211, p= 0.008), BRC funding (U=5335, p=0.023), external grant funding, (U=5444, p=0.042), collaboration with industry (U=5434, p=0.041), organisational policies on gender equity (U=5462, p 0.034), and organisational targets (U=5375, p=0.026 (Table 2).

Overall, all 13 markers of GE were ranked as the most important marker by a higher proportion of female participants compared with the male participants (Fig 2).

**Fig 2.**
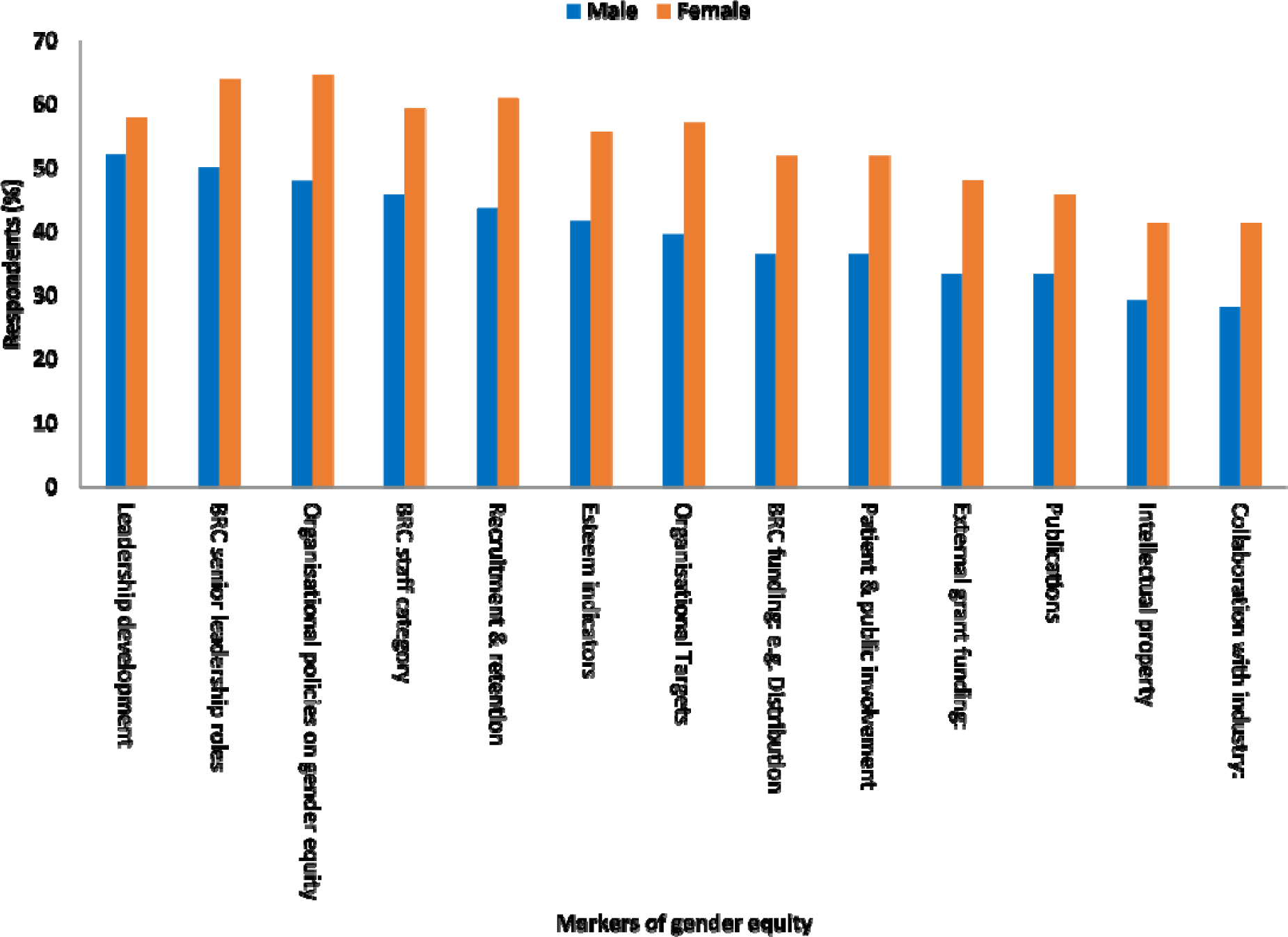
Markers of gender equity marked as very important by gender.

We created a priority ranking order from 1 to 13 of all markers of GE based on the highest (Rank 1) and the lowest (Rank 13) percentage of participants ranking each marker as very important (Table 3).

**Table 3.**
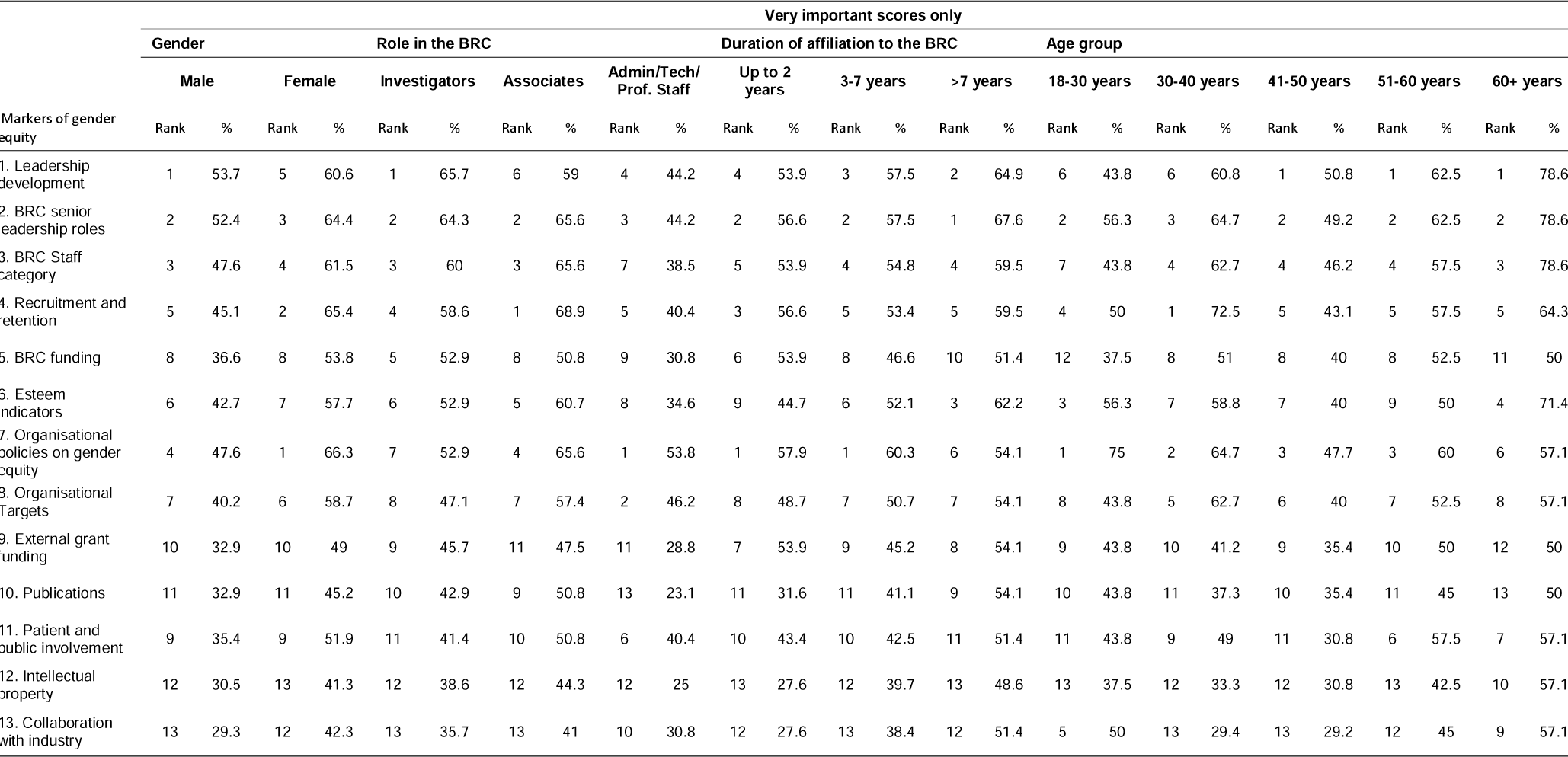
Priority ranking order of markers of gender equity by participants’ gender, role in the BRC, duration of affiliation with the BRC and age.

Analysis of ranking by gender of participants showed that the top two markers of GE from the male participants’ perspective were “leadership development” and “BRC senior leadership roles”, which were ranked as very important by 53.7% and 52.4% of male participants. Conversely, “organisational policies on gender equity” and “recruitment and retention” were the top two most important markers of GE ranked by 66.3% and 65.4% of female participants (Table 3).

Results showed that the top most important marker of GE in BRC was different for different categories of participants’ roles in the BRC. “Leadership development” was the most important marker for 65.7% of investigators, “recruitment and retention” for 68.9% of associates and “organisational policies on gender equity” for 53.8% of admin, technical and professional staff (Table 3). The Kruskal-Wallis H test showed that the mean scores of ranking the importance of the markers by the role in the BRC were statistically significantly different for eight markers: BRC senior leadership roles (χ^2^ (2) = 8.89, p = .012), BRC leadership development (χ^2^ (2) = 6.98, p = .031), BRC staff category (χ^2^ (2) = 14.39, p = .001), recruitment and retention (χ^2^ (2) = 13.25, p = .001), BRC funding (χ^2^ (2) = 11.46, p = .003), external grant funding (χ^2^ (2) = 12.12, p = .002), esteem indicators (χ^2^ (2) = 7.1, p = .029) and publications (χ^2^ (2) = 10.59, p = .005) (Table 3).

Analysis of rankings by the duration of affiliation to the BRC identified that “BRC senior leadership roles” was the most important marker of GE for 67.6% of participants who had worked for 7 years or more while “organisational policies on gender equity” was the most important marker of GE for 57.9% and 60.3% of participants who had been affiliated to the BRC for up to 2 years and 3-7 years, respectively (Table 3).

Ranking by the age of participant demonstrated that “Leadership development” was the top most important marker of GE for participants aged 40 years and more while the “Organisational policies on gender equity” was the most important marker for 75% of the youngest participants (aged 18-30 years) and “recruitment and retention” for 72.5% of participants who were 30-40 years old (Table 3). However, Kruskal-Wallis H tests showed that there were no statistically significant differences in the mean rankings of all markers between different categories of participants’ age and duration of affiliation to the BRC.

### Exploratory factor analysis

Results of the first EFA model that included all 13 measured markers revealed a two factor solution but the rotated component matrix showed that marker No. 5 (i.e., BRC funding) had very high cross loadings i.e., 0.53 on factor 1 and .68 on factor 2. We removed this marker and re-ran the EFA model with 12 markers, which again resulted in a 2 latent factor solution but again the rotated component matrix revealed that measured marker No. 7 (i.e., esteem indicators) had very high cross loadings i.e., 0.50 and 0.67 on factor 1 and factor 2 respectively. Consequently, we removed marker No. 7 and re-ran the EFA, which again resulted in a 2 latent factor solution with the rotated component matrix showing marker No. 11 (i.e., patient and public involvement) with higher cross loadings i.e., .61 on factor 1 and .45 on factor 2.

Subsequently, we removed marker No. 11 from the EFA and re-ran the model, which showed no marker having cross loading ≥0.45 on more than one factor “Table 4”. We selected this model as a final EFA model with the Kaiser-Meyer-Olkin (KMO) Measure of Sampling Adequacy = .884 and statistically significant Bartlett’s Test of Sphericity (χ^2^ = 1553.69, p < 0.001), which confirmed the suitability of the data for running the EFA model. Table 4 presents the statistics about the extracted communalities, total variance explained and Rotated Factor Matrix. Based on the content of loaded measured items on latent factor 1 and factor 2, we identified these markers as the organisational and individual markers respectively (Table 4).

**Table 4.** Exploratory factor analysis: Measured markers of gender equity, latent factors with loadings, communalities, Eigen values, KMO Measure of Sampling Adequacy, Bartlett’s Test of Sphericity, total variance explained and scale reliabilities.

| Measured items/ Markers of gender equity | Rotated Component Matrix<sup>a</sup> |  |  |
| --- | --- | --- | --- |
|  | Component loadings |  | Communalities |
| | Factor 1<br>(Organisational markers) | Factor 2<br>(Individual markers) | $h^2$ |
| BRC senior leadership roles | **.84** | .30 | .80 |
| Leadership development | **.81** | .29 | .74 |
| BRC staff category | **.78** | .39 | .76 |
| Recruitment & retention | **.77** | .28 | .67 |
| External grant funding | .42 | **.79** | .79 |
| Publications | .34 | **.83** | .80 |
| Intellectual property | .34 | **.87** | .87 |
| Collaboration with industry | .30 | **.85** | .81 |
| Organisational policies on gender equity | **.68** | .33 | .58 |
| Organisational Targets | **.72** | .35 | .64 |
| *Eigenvalues* | 6.40 | 1.06 |  |
| *Kaiser-Meyer-Olkin (KMO) Measure of Sampling Adequacy* | .884 |  |  |
| *Bartlett's Test of Sphericity* | $\chi^2 = 1553.69$ | | |
| *Significance (P)* | < 0.001 |  |  |
| *of total variance explained* | 63.98 | 10.65 |  |
| *Cronbach's $\alpha$ reliability (Standardised)* | .912 | .925 |  |

<sup>a</sup>Extraction Method: Principal Component Analysis; Rotation Method: Varimax with Kaiser Normalization; Rotation converged in 3 iterations.

### Confirmatory factor analysis

To check the two latent factor solution observed in the EFA (Table 4), we ran CFA (henceforth mentioned as the initial CFA model) as shown in Fig 3A.

**Fig 3A.**
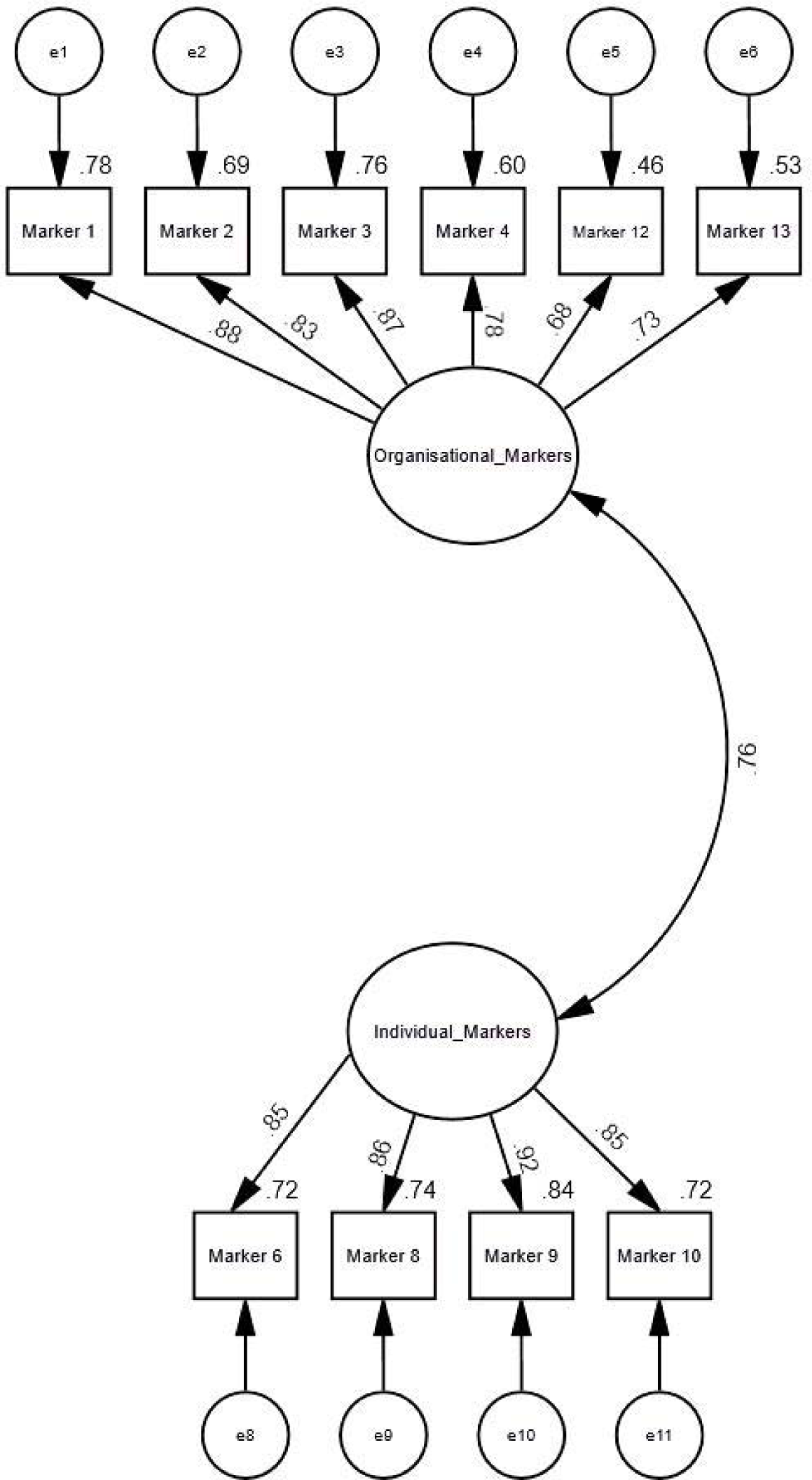
Initial CFA-model. **Notes:** Rectangles represent measured items (endogenous variables); circles represent latent (unmeasured/exogenous) variables. Marker 1 (BRC senior leadership roles), Marker 2 (Leadership development), Marker 3 (BRC staff category), Marker 4 (Recruitment and retention), Marker 6 (External grant funding), Marker 8 (Publications e.g. authorship), Marker 9 (Intellectual property), Marker 10 (Collaboration with industry), Marker 12 (Organisational policies on gender equity) and Marker 13 (Organisational Targets for gender equity).

A model summary of goodness of fit (GoF) indices for the initial CFA model is presented in (Table 5). The GOF indices suggested that the initial CFA model did not fit with the data. Consequently we applied some of the modifications indices suggested in the results. This led to addition of a few correlations between error estimates of some measured items as shown in Fig 3B (henceforth mentioned as the post-hoc CFA model).

**Fig 3B.**
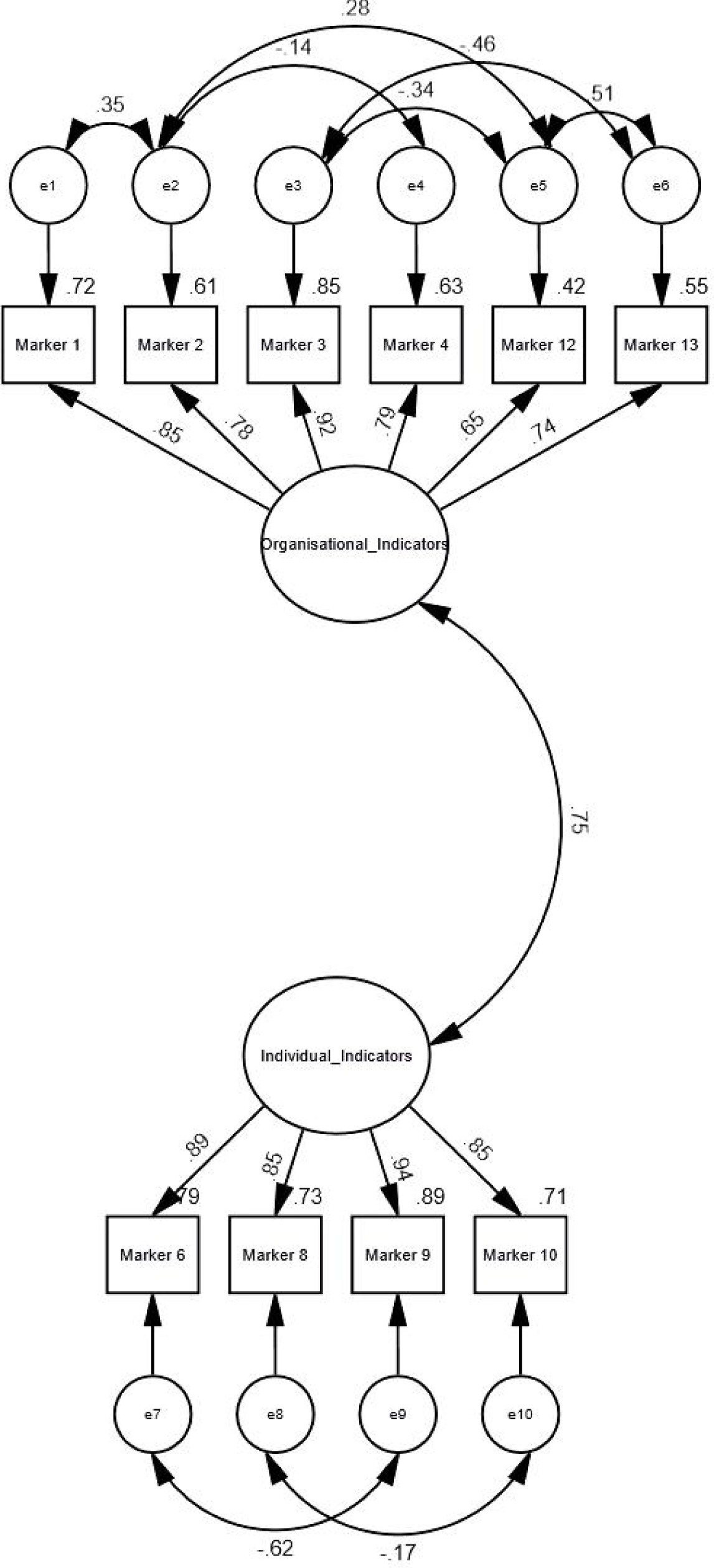
Post-hoc CFA model. **Notes:** Rectangles represent measured items (endogenous variables); circles represent latent (unmeasured/exogenous) variables. Marker 1 (BRC senior leadership roles), Marker 2 (Leadership development), Marker 3 (BRC staff category), Marker 4 (Recruitment and retention), Marker 6 (External grant funding), Marker 8 (Publications e.g. authorship), Marker 9 (Intellectual property), Marker 10 (Collaboration with industry), Marker 12 (Organisational policies on gender equity) and Marker 13 (Organisational Targets for gender equity).

**Table 5.** Summary of goodness of fit indices observed in the initial and post-hoc CFA models.

| Values | $\chi^2$ | Df | Sig (p) | $\chi^2/df$ | IFI | NFI | CFI | RMSEA |
| --- | --- | --- | --- | --- | --- | --- | --- | --- |
| Recommended | | | >0.05 | $\leq 3.00$ | $\geq 0.9$ | $\geq 0.9$ | $\geq 0.9$ | $\leq 0.08$ (p>0.05) |
| Observed in initial CFA model | 239.95 | 43 | <0.001 | 5.58 | 0.88 | 0.86 | 0.88 | 0.16‡ |
| Observed in Post-hoc CFA model | 48.81 | 26 | 0.004 | 1.88 | 0.98 | 0.97 | 0.99 | .07‡(Low 90.038, High 90.098), p Close 0.141 |

$\chi^2$ , Chi-square; Df, degrees of freedom; Sig, significance level (p), CFI, comparative fit index; IFI, Incremental fit index; NFI, normed fit index; RMSEA, root mean square error of approximation; ‡90 confidence interval for RMSEA

The GOF indices of the post-hoc model (Table 5) showed a good fit of the post-hoc model with the data. We therefore accepted the post-hoc CFA model as the final CFA model.

The estimates of standardized regression weights (β) of measured items on to the latent factors along with their significance level (p) observed in both the initial and the post-hoc CFA models (Table 6), demonstrate that all measured markers had statistically significant higher loadings on organisational makers factor (β ≥0.68, p<0.001) and individual markers factor (β ≥0.85, p<0.001) (Table 6, Figs 3A and 3B). We checked the internal consistency and convergence of both latent factors by the composite reliability and average variance extracted (AVE) respectively. Six measured items (markers) that loaded on to the organisational markers factor explained 64 and 63 AVE in the initial and the post-hoc CFA models respectively whereas four items that loaded on to the individual markers factor explained 76 and 78 AVE in the initial and post-hoc CFA models respectively. The observed AVE for both factors was higher than the minimum 0.5 that is suggested for adequate convergence [17,18].

**Table 6.** Latent factors, measured markers and standardised estimates observed in initial and post-hoc CFA models.

| Latent factors | Measured markers | Initial CFA Model |  |  | Post-hoc CFA Model |  |  |
| --- | --- | --- | --- | --- | --- | --- | --- |
|  |  | Estimate†<br>(β) | C.R. | P | Estimate†<br>(β) | C.R. | P |
| Organisational Markers | → Marker 1 | 0.88 | ‡ | ‡ | 0.85 | ‡ | ‡ |
|  | → Marker 2 | 0.83 | 15 | *** | 0.78 | 15.9 | *** |
|  | → Marker 3 | 0.87 | 16.4 | *** | 0.92 | 16.1 | *** |
|  | → Marker 4 | 0.78 | 13.3 | *** | 0.79 | 13.2 | *** |
|  | → Marker 12 | 0.68 | 10.7 | *** | 0.65 | 9.34 | *** |
|  | → Marker 13 | 0.73 | 12.1 | *** | 0.74 | 10.3 | *** |
| Individual Markers | → Marker 10 | 0.85 | ‡ | ‡ | 0.85 | ‡ | ‡ |
|  | → Marker 9 | 0.92 | 16.7 | *** | 0.94 | 16.1 | *** |
|  | → Marker 8 | 0.86 | 15.1 | *** | 0.86 | 14.1 | *** |
|  | → Marker 6 | 0.85 | 14.7 | *** | 0.89 | 14.5 | *** |

Marker 1 (BRC senior leadership roles), Marker 2 (Leadership development), Marker 3 (BRC staff category), Marker 4 (Recruitment and retention), Marker 6 (External grant funding), Marker 8 (Publications e.g. authorship), Marker 9 (Intellectual property), Marker 10 (Collaboration with industry), Marker 12 (Organisational policies on gender equity) and Marker 13 (Organisational Targets for gender equity), †Estimates of standardized regression weights, ‡Not estimated because of loading set to fixed value i.e., 1.0, \*\*\* Significance value (p): < .001

We calculated the composite reliability for the organisational markers factor as 0.91 for both the initial and the post-hoc CFA models and the composite reliability for the personal markers factor as 0.93 and 0.94 in the initial and post-hoc CFA models respectively. The composite reliabilities for both latent factors were higher than the minimum required composite reliability of 0.7, which indicated that both latent factors have a high internal consistency suggesting that the loaded measured markers (items) consistently represented the respective identified latent factor [17,18]. The CFA models showed that both latent factors i.e., the organisational markers and the individual markers have a strong correlation i.e., 0.76 and 0.75 in the initial and post-hoc CFA models respectively. The EFA and CFA results identified and confirmed two significant dimensions i.e., organisational and individual markers of GE in BRCs.

### Analysis of open ended comments

Participants were asked two open ended questions: “Please state any other indicators related to gender equity that the BRC should assess and monitor from the questionnaire?” and “Provide any comments or suggestions on new ways of measuring gender equity in Biomedical Research Centres”. Sixty eight (28%) participants provided open ended comments. Twenty nine participants suggested other indicators related to gender equity (20 female, 6 male, 2 prefer not to say and 1 self-describe). Thirty nine participants provided open ended comments on new ways of measuring gender (24 female, 14 male and 1 self-describe). Participants’ open ended comments identified three key themes. The most frequent comments concerned ‘monitoring and benchmarking of gender’ followed by ‘organisational policies to support childcare responsibilities’ and ‘leadership and Institutional support’, as described below.

#### Monitoring and benchmarking gender

Actions to monitor and benchmark gender at an organisational level were the most frequent response. Many respondents suggested a range of areas to monitor by Gender: occupational role and seniority, gender balance of speakers in scientific sessions, allocation of funding, career development and recruitment and retention. However the complexity of benchmarking gender was raised, additional factors such as part time working, differing gender identities and relative numbers of women working full time were important.

> “*I think analysis by role is important - you can see that particular roles are heavily filled by one gender. Why this is occurring needs to be understood and addressed*.*”* (R50, Male)
>
> *“For any BRC post or call for funding scheme, please publish how many male/female applicants were received, and how many male/female achieved the post/ward…merit/track record should be the primary assessment criterion, it would also be concerning if only one sex/gender tends to predominate the winners list. Equally, one should not fill a post or award a grant just based on sex/gender if the applicant is not qualified - this creates resentment and undermines the effort that aims to genuinely try to provide equal opportunities to both sexes (or if we are talking about gender, all the different categories of genders)*.*”* (R273, Female)
>
> “*It is very important that measures of gender equity are appropriately related to numbers of FTE staff i*.*e. female researchers may have fewer outputs on average, but this should clearly be interpreted in relation to the correct denominator (i*.*e. FTE) in order to account for part-time working*.*”* (R213, Male).

### Career development

Respondents proposed a range of actions from benchmarking the career development of female early career researchers and overall career progression within the BRC.

> *“Analysis of career progression for women as early career researchers to senior positions required*.*”* (R153, Female)
>
> *“Career progression since first significant association with the Oxford BRC*.*”* (R32, Female)

### Recruitment and retention

Participants felt it was important to monitor recruitment and retention for GE and proposed a range of actions. These included monitoring the seniority of staff and gender to assist exploring retention and recruitment processes, gender balance of interview panels and benchmarking number of applicants for posts by gender.

> *“Recruitment and retention, alongwith career development should be closely monitored for gender distribution*.*”* (R68, Female)
>
> *“Compare number of senior investigators and Professors with the numbers and sex of postdocs and doctoral students, where are people dropping out? or where are the recruitment practices potentially biased?”* (R227, Male) *“Interview panels for new staff*.*”* (R50, Male)

### Organisational support for those with childcare responsibilities

The second most frequent actions concerned organisational support for those with childcare responsibilities. Many respondents proposed introducing specific funds to support maternity leave, childcare costs and caring responsibilities. Changing the timings of departmental events and meetings to facilitate attendance of those with childcare responsibilities was raised by several respondents.

> **“***A research enablement fund for researchers with caring responsibilities would be very helpful*.*”* (R246, Female)
>
> “*Provide greater support to women to be able to achieve the same as men e*.*g. childcare support costs while attending meetings and conferences*.*”* (R228, Female)
>
> “*Participation in departmental seminars / workshops: often these are timed to go on beyond the end of the working day, which excludes anyone with childcare responsibilities (predominantly women/early career researchers) from fully participating*.*”* (R228, Female)
>
> *“Status as a carer - e*.*g. primary carer (so that gender roles in supporting partners can be monitored - for example, men/women taking primary carer/dual carer roles to support women/men going back to work)”*. (R270, Male)

### Leadership and Institutional support for Gender equity

Respondents raised the importance of specific policies at an institutional level to support GE. Some respondents stated that ensuring gender diversity in senior leadership roles would positively impact on the rest of the organisation. Others described how GE equity policies should be in place for all genders in the organisation. Inequity in pay was also raised by participants as an important marker of GE.

> “*Policy on promoting women in science and at an HR level required*.*” (R153, Female)*
>
> *“If ‘influencer’ roles are appropriately gender-diverse, then policies, training programmes etc. will reflect that”*. (R129, Prefer not to state gender)
>
> *“My personal view is that it is ensuring the policies and opportunities are there for both genders throughout the organisations that is the most important thing”*. (R125, Male)
>
> *“Equity in pay*.*”* (R172, Female)

## Discussion

### New statistically significant model of gender equity markers

Our survey identified a new statistically significant model of GE markers with two distinct dimensions of GE markers, i.e. organisational markers and individual markers (Fig 4), which are discussed below. The present study is the first, to our knowledge, that has developed such a model.

**Fig 4.**
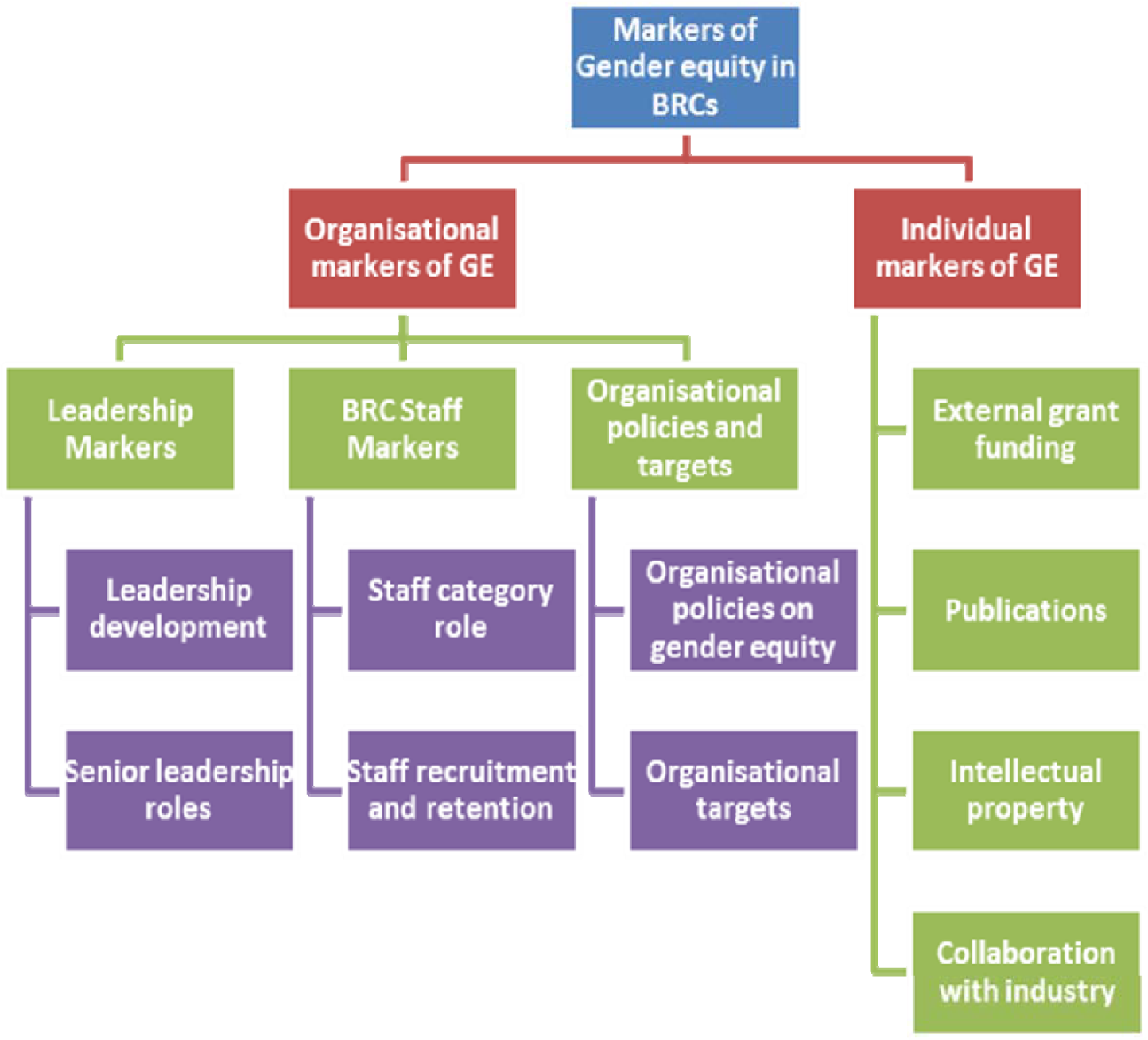
Organisational and individual markers of gender equity in BRCs.

The response rate (36%) in our study is consistent with online questionnaire survey response rates which are typically lower than mail based questionnaires [24] and when participants include clinical professionals [25]. Our results show that the majority of respondents were female suggesting that women were slightly more likely to respond. Research has shown that the relevance of the study topic may impact response rates and so this may have been a factor too [26]. The results also show that women and men rank the importance of the markers of GE differently and a greater proportion of women ranked all markers as the most important compared to the proportion of male participants. However a relatively high proportion of respondents were male (40) this is key as research has indicated men’s support and perspective is also an important driver of GE in institutions [27].

### Leadership roles

When scores for all participants were combined, BRC senior leadership roles ranked as the most important marker of GE followed by organisational policies on gender equity (Fig 4) This may reflect findings from a recent evaluation of the Athena SWAN programme which highlighted significant challenges remain in addressing gender balance in the most senior positions in higher education (e.g. professorial, senior management) [9]. It also supports the finding that leadership is a key driver for sustainable organisational change in terms of GE [12].

### Organisational policies on gender equity

Our results suggest that organisational policies on GE are an important measure required. Organisational policies also emerged as a key theme in the open ended comments where a range of specific actions were identified underlining the benefits of open ended questions to provide detailed illustrations of the responses to the closed questions [29]. This is typically facilitated at an organisational level by Athena Swan but the results suggest more BRC focussed targets may be beneficial at a local level. Linking the direct impact of the introduction of Athena SWAN to the acceleration of GE in an institution is challenging due to the complexity of issues [12].

### Analysis of ranking by gender of participants

Analysis of ranking by gender of participants showed that the top two markers of GE from the male participants’ perspective were “leadership development” and “BRC senior leadership roles”, which were ranked as very important by male participants. Conversely “organisational policies on gender equity” and “recruitment and retention” were the top two most important markers of GE ranked by 66.3 and 65.4 of female participants (Table 3).

### Markers of achievement in industry and gender equity

Interestingly, collaboration with industry and the Intellectual property emerged as the last and second last very important markers of GE in BRCs reported by 35.4 (n=86) and 35.8 (n=87) of participants respectively (Table 2). This may reflect relatively low participation of women in industry [28]. However it is an important area to address given that collaboration with industry is an important metric in BRCs to report to their funder the NIHR.

### Organisational markers of gender equity

The dimension of organisational markers of GE identified in our study comprised six markers of GE (Tables 4 and 6, Fig 4). The scrutiny of the wording and content of these six markers by authors showed these six markers could be sub-divided into three sub-dimensions; leadership markers, BRC staff markers and organisational policies and targets markers (Fig 4).

#### Leadership

The leadership sub-dimension comprised two markers i.e., leadership development and senior leadership roles. The BRC staff sub-dimension included two markers i.e., staff category and staff recruitment and retention. While the third sub-dimension of organisational policies and targets encompassed two markers: organisational policies on gender equity and organisational targets (Fig 4).

The high ranking of leadership for GE in this study suggests further local organisational policies may be appropriate. As highlighted elsewhere local drivers are important to support existing GE initiatives for example a recent evaluation of Athena SWAN did not indicate a statistical relationship between the Charter and increase in the proportion of female staff over time [10]. The open ended questions also supported the findings that leadership and institutional support are important drivers for GE.

#### Organisational policies

Our study provides new potential GE markers within the specific setting of NIHR BRCs. The population is intentionally broader than previous GE research which has predominantly focussed on clinical academic settings [30–32] or Athena SWAN research project populations which focus on university and academic staff [10,11]. In contrast this study population is intentionally broader including both NHS staff and clinical and non-clinical university staff at all levels.

### Individual markers of gender equity

The second dimension i.e., individual markers of gender equity identified included four markers of GE (Tables 4 and 6, Fig 4). The review of the wording and content of these four markers by authors suggested these four markers could be sub-divided into four sub-dimensions, which include research funding, publications, intellectual property and industry collaboration and each of these sub-dimensions included only one GE marker i.e., marker 6, 8, 9 and 10 respectively (Tables 4 and 6). These findings concur with a recent analysis of lessons learned from the Athena SWAN demonstrates the importance of baseline data for the purposes of benchmarking, and importance of leadership to enable systemic change [12]. At the NIHR Oxford BRC, benchmarking of gender and BRC publications and staff is in place. However, TRO funders may consider encouraging gender benchmarking or making it mandatory. For example, currently the only mandatory request for gender data within BRCs is NIHR academy members.

### Strengths and limitations

To our knowledge this is the first study to explore views on new markers of achievement for women in academic science specifically in an NIHR BRC. Previous research in this field has focussed predominantly on clinical academic settings and Athena Swan evaluations [11,12,32]. This study has a broader remit to develop new ways of monitoring gender equity. This study extends understanding of GE as set out in our research protocol to understand “new ways of assessing and monitoring gender equity in translational research organisations”[14].

Our two factor model of gender equity markers (Fig 4) extends the Multidimensional conceptual framework for gender equity assessment and monitoring [15] to address the gap in GE in TROs as it illustrates the prioritisation of markers of gender equity based on BRC affiliates perspectives in an NIHR BRC. It also examines both women’s and men’s perceptions to enable gender comparative research and the response of men was relatively high (40%). It is important to include men in such surveys if changes at an organisational level are to be sustainable and relevant to all and to ask the same questions to enable gender comparative research [26]. Given the significant investment in NIHR BRCs and direct link of demonstrable progress in GE equity this is of particular relevance to policy makers [6]. A limitation of the current study is that we did not collect demographic information on ethnicity this could be adopted in future studies.

It is important to acknowledge measuring GE equity is complex given the differing identities of women and men. Furthermore gender has been defined as culturally defined social constructs associated with being female or male [36]. Due to the relative low numbers we removed the category “self-identify” from the final analysis. Future research should explicitly take this diversity into account when planning online surveys. It is also important to avoid gender stereotypes and biases [37].

This questionnaire survey provides important insights into views and measures on Gender equity however GE is complex and there are a number of factors such culture, leadership, bias and transformational change which take significant time and are difficult to measure and may influence the generalisability of the findings (12,13,32).

Evidence suggests that culture in academic medicine may impact on career progression for women [32,38]. Whilst this study did not examine culture specifically such issues were raised in the open ended comments of the questionnaire. As other research has highlighted there are external factors which need to be reviewed to enact long term structural change such as “cultural change” and the role of men in work life balance [11]. Benchmarking data is also important however there are external cultural and societal norms and biases that need to be taken into account and were out with the scope of this survey. Such issues were again raised in the open ended comments of the questionnaire. The open ended comments identified three key areas of action which are beneficial to the development of future work in this field and are in alignment with Athena SWAN which proposes specific action plans are developed and monitored [7]. Given the topic of GE and self-selection of participants to respond they may reflect strong views on GE not be generalizable to other settings. However they do offer the opportunity to provide more detailed responses to the closed question responses [29]. This survey includes men’s perspectives in addition to women’s perspectives on GE, which is important because for change to be sustainable men’s input and perspective is important. This survey contributes to the evidence base on gender equity in a translational research organisation. High quality data is essential for measuring institutional change in addition to baseline data for benchmarking purposes [12].

## Conclusions

Our findings have important implications to inform prospective planning and monitoring within the field of organisational policies and leadership policies to accelerate women’s advancement and leadership within NIHR BRCs. Research has highlighted GE in the workforce is an important indicator for internationally competitive organisations. Implementation and sustainability of gender equity however requires strong commitment at senior leadership and organisational policy level. We advocate for enhanced collaborations across NIHR biomedical research centres (currently n=20) in the field of GE to provide larger data sets which can inform broader understanding of progress but also barriers to acceleration in the GE domain.

## Data Availability

All data are reported in the manuscript

## Acknowledgments

The authors wish to thank the participants for taking part in this study and completing the survey.

